# The LIBERATE Internal Randomized Controlled Pilot Trial: Adjunctive midodrine therapy for early liberation from vasopressor support in critically ill patients

**DOI:** 10.1101/2025.07.31.25332174

**Authors:** Dawn Opgenorth, Fadi Hammal, Deborah J. Cook, D’Arcy Duquette, Stephanie Sibley, Jennifer Tsang, Janek Senaratne, Kirsten Fiest, Vincent I Lau, Fernando G Zampieri, Sean M Bagshaw, Oleksa G Rewa

**Affiliations:** Department of Critical Care Medicine, Faculty of Medicine and Dentistry, University of Alberta, and Alberta Health Services, Edmonton AB, Canada; Department of Medicine, McMaster University, McMaster Health Sciences Center, Hamilton, ON, Canada; Critical Care Program Improvement and Integration Network, Alberta Health Services, Alberta, Canada; Department of Critical Care Medicine, Queens University, Kingston, ON, Canada; Niagara Health Knowledge Institute, Niagara Health, St Catharines, ON, Canada; Cumming School of Medicine, University of Calgary, Calgary, AB, Canada

**Keywords:** Intensive Care Unit, Critical Care, Shock, Vasopressors, Midodrine

## Abstract

**Background:** Intravenous (IV) vasopressors are the mainstay of hemodynamic physiological support for critically ill patients. However, the role of oral vasopressors in the management of IV vasopressor-dependent shock remains unclear. The LIBERATE body of work aims to examine the role of midodrine as an IV vasopressor sparing agent for critically ill patients with IV vasopressor dependent shock. The objective of the LIBERATE internal randomized controlled trial was to evaluate the implementation of the LIBERATE protocol in a multi-centre randomized control trial setting.

**Methods:** We conducted a pilot multi-centre, concealed-allocation, parallel-group, blinded randomized controlled trial (RCT) evaluating the effect of midodrine 10mg q8h versus placebo on the duration of IV vasopressor-dependent shock in the intensive care unit (ICU) on the first 200 of planned sample size for the LIBERATE trial. The study was performed in 9 Alberta and Ontario medical centres between September 16, 2022 and January 22, 2025. We included patients aged 18 years or older admitted to the ICU receiving ongoing IV vasopressor support. Patients were randomly assigned 1:1 to enteral midodrine or an identical placebo and received the study treatment until 24 hours after discontinuation of their IV vasopressor therapy. The primary endpoints were study feasibility and included: recruitment rate (monthly enrollment per study site), screening and enrollment characteristics, protocol adherence, retention rate, data completeness, and safety (adverse event reporting), Secondary endpoints focused on aggregate patient-centered outcomes.

**Results:** There were 200 patients who fulfilled eligibility criteria and were enrolled in the trial. The mean age was 61.7 (SD 15.0), and 80 (40.1%) were female. The mean Acute Physiology and Health Evaluation (APACHE II) score at ICU admission was 23.7 (SD 7.6) and the mean Sequential Organ Failure Assessment (SOFA) score at the time of study enrollment was 10.1 (SD 2.9). Sepsis was the most common cause of shock in both groups. Average recruitment was 1.4 participants (range 0.3 to 3.0) per study site per month, protocol adherence was 84.5%, retention rate was 97.0%, and there were no reports of unblinding of treatment allocation by the study sites. Two adverse events were reported and resolved with no sequelae.

**Conclusion:** The LIBERATE internal randomized controlled trial is feasible and safe, supporting a larger trial to investigate the utility of midodrine in critically ill patients with IV vasopressor-dependent shock.

**Trial Registration:** Registered September 28, 2021. https://clinicaltrials.gov/study/ NCT05058612

**Key messages regarding feasibility:**

- It was previously unknown if recruitment rates, protocol adherence and safety of midodrine would remain in effect across multiple centres
- This internal randomized controlled trial across 9 sites established sufficient feasibility outcomes with recruitment rates of 1.4 participants per site per month, protocol adherence of 84.5%, retention rates of 97.0%, data completeness of 96.0% and adverse event rate attributable to study medication of 0%.
- The results of this trial will be used to inform and proceed with the LIBERATE RCT.

## Background

Intravenous (IV) vasopressors are the standard of care for critically ill patients who develop shock.[1–5] This is necessary in 15-40% of critically ill patients.[1] However, vasopressors for hemodynamic support can be associated with risk of tissue necrosis, vascular injury, cardiac dysrhythmias, and bowel ischemia.[6] Oral vasopressors do not carry these risks.[7] Current clinical practice guidelines do not include a statement on the use of oral vasopressor therapy, and only provide support for the use of IV vasopressors for hemodynamic management of critically ill patients in shock.[8] An effective oral vasopressor could plausibly shorten the duration of IV vasopressors, reduce the risk of adverse effects, and allow earlier transition of patients from the intensive care unit (ICU).

Midodrine hydrochloride is a commonly used alpha-agonist oral vasopressor, and has been traditionally used to reduce symptoms of orthostatic and intra-dialytic hypotension in non-critically ill patients.[9–12] Midodrine has good oral bioavailability (i.e., 93%), and a half-life of 3 hours, allowing less frequent dosing and auto-off titration when stopping the medication. A systematic review by our study team identified 17 studies evaluating the role of midodrine in 2,567 critically ill patients.[13] In pooled analysis from 6 RCTs published in complete form (i.e., 374 patients), we found that midodrine may decrease ICU length of stay (LOS) by 1.01 days when compared with placebo (low certainty of evidence). Findings of this systematic review provided strong rationale for a more robust and rigorous evidence base to understand the role for midodrine in critically ill patients receiving IV vasopressors.[14–19]

The Adjunctive midodrine therapy for early liberation from vasopressor support in critically ill patients (LIBERATE) internal randomized controlled trial is a multi-centre, concealed-allocation, parallel-group, blinded randomized controlled trial (RCT) that tests the hypothesis that use of adjunctive midodrine will decrease ICU LOS in ICU patients with IV vasopressor–dependent shock. Previously, we conducted a single-centre RCT to refine the study protocol demonstrate the feasibility and safety of the trial design.[20] As a next step we have conducted a multi-centre pilot RCT to assess the feasibility of timely recruitment and protocol adherence in the implementation of the protocol in a multi-centre setting to include both academic and community sites. The results of the LIBERATE internal randomized controlled trial will inform the LIBERATE RCT.

## Methods

### Study Design

We conducted an internal pilot multi-centre, concealed-allocation, parallel-group, blinded randomized controlled trial (RCT) in 9 Canadian mixed medical surgical ICUs, in which patients were allocated to a placebo or midodrine in a 1:1 ratio. We included the first 200 patients from the sample size of the planned LIBERATE RCT to ensure sufficient sample size. Outcome data will not be unblinded as accumulated data will be utilized in the main analysis set. A person with lived experience assisted with the design of this research.

This study was registered with clinicaltrials.gov on September 28, 2021 (NCT05058612).

### Ethics and consent for participation

This study was approved by the University of Alberta Research Ethics Board (Alberta Research Information Services (ARISE)) on November 26, 2021 (Pro00112293) and the Queen’s University Health Sciences and Affiliated Teaching Hospitals Research Ethics Board on September 1, 2023 (Clinical Trials Ontario Project ID: 4459).

Informed consent was provided by either the patient or their surrogate decision maker (SDM). If the patient lacked capacity and an SDM could not be identified or contacted at the time of study eligibility, a deferred consent model was utilized (also known as consent to continue), and written consent was obtained from the patient once they regained capacity. If participants did not provide consent post regaining capacity, they were excluded. There is no a priori plan to provide participants with the results of this study.

### Study setting

The trial was conducted at 3 academic and 6 community hospitals in Alberta and Ontario. The Research Office in the Department of Critical Care Medicine at the University of Alberta was the Coordinating Centre and responsible for protocol development, data capture, preparation and dispensing of the study investigational product (IP) and the overall coordination and maintenance of study operations. The study was monitored by the Clinical Trials Office (CTO) at the University of Alberta (https://www.ualberta.ca/en/health-sciences/research/university-of-alberta-clinical-trials/clinical-trials-office.html). Further information regarding the LIBERATE study is available at https://www.ualberta.ca/en/critical-care/research/liberate/index.html and the LIBERATE RCT is registered with ClinicalTrials.gov (NCT05058612).

### Inclusion criteria

1. Adult patients aged ≥18 years
2. Admitted to ICU
3. Receiving a continuous infusion of IV vasopressor support, defined by a minimum of any one of norepinephrine ≥0.05 mcg/kg/min, epinephrine ≥0.05 mcg/kg/min, vasopressin ≥0.04 U/min, or phenylephrine ≥0.1mcg/kg/min
4. Evidence of a decreasing vasopressor dose, defined as the dose at the time of screening being less than peak dose in the preceding 24 hours were screened for eligibility.

### Exclusion criteria

1. Over 24 hours from peak vasopressor dose
2. Contraindication to enteral medications
3. Known allergy to midodrine
4. Had received midodrine in the last 7 days prior to screening
5. Were expected to die or have withdrawal of life-sustaining therapies in the next 24 hours
6. Known pregnancy.

### Study intervention

All participants received standardized vital and physiological monitoring while they received the study intervention. Monitoring included continuous heart rate, 3-lead electrocardiogram (ECG), pulse oximetry and intravascular blood pressure monitoring. Vital signs and physiological parameters including but not limited to respiratory rate, non-invasive blood pressure monitoring, Glasgow Coma Scale, and urine output were monitored hourly – or more frequently – as needed. Patients randomized to the experimental treatment received midodrine 10 mg administered enterally, every 8 hours from time of study inclusion to 24 hours after IV vasopressor termination. This dose was chosen as it was the most commonly used dose of midodrine used in previous studies and the highest dose as per the product monograph.[21] Due to its prolonged half-life when compared to norepinephrine, midodrine does not require a tapering of dose.

Patients randomized to the control arm received a placebo administered enterally every 8 hours from time of study inclusion to 24 hours after IV vasopressor termination. The study investigational product (IP) was prepared by a compounding pharmacy in Edmonton, Alberta (https://www.strathconapharmacy.com/strathcona-compounding-ltd-.html) and underwent dosage testing at an accredited laboratory (https://www.keystonelabs.ca/) prior to dispensation. The IP was prepared, labelled and dispensed by an unblinded research pharmacy and administered by the blinded bedside nurse. If the patient had an oral/nasal feeding tube, the contents were removed from the study capsule, mixed with water and administered through the feeding tube.

Sites without a dedicated research pharmacy received pre-dispensed, blinded study drug containers prepared by the Edmonton compounding pharmacy, labeled with randomization codes and assigned sequentially upon patient enrollment.

### Randomization, blinding and allocation concealment

Patients were randomized in a 1:1 ratio to midodrine or placebo using permuted blocks of undisclosed and random variable size. Randomization was stratified by site.

Intensivists, research personnel, patients, the ICU treating team, members of the executive and steering committees and data analysts were blinded to the treatment allocations. The midodrine and placebo IP were indistinguishable, in identical capsule form. Only the research pharmacy team responsible for conducting the treatment allocation and preparation of the IP were unblinded to prepare study drug.

### Primary feasibility outcomes

The primary feasibility outcomes were, 1) recruitment rates, with a target average of ≥ 1 patient per site per month; 2) screening and enrollment characteristics; 3) protocol adherence for administered study medication, with a target protocol adherence of ≥ 80%; 4) retention rate, with a target of having ≥ 90% of recruited participants completing study outcomes, 5) data completeness, with a target of ≥ 95% of all data being collected as intended; and 6) adverse event, with a target of monitoring and reporting on all adverse events. If feasibility criteria were not met, then progression to full trial would not occur.

### Secondary clinical outcomes

The secondary clinical outcome measures included: ICU mortality, ICU and hospital length of stay, duration of IV vasopressor support, 90-day all-cause mortality, ICU readmission, and re-initiation of IV vasopressors.

### Data collection

Data were collected from the patient clinical record and entered into an electronic case report form (CRF) (REDCap, Vanderbilt [https://project-redcap.org/]). Study sites were provided with an identical paper CRF as a data collection tool; however, study coordinators could opt to enter data directly into the electronic CRF, and only electronic CRF data was reviewed by the study monitor. Variables collected included demographics, baseline characteristics, daily ICU treatments (mechanical ventilation, renal replacement therapy, vasopressors, corticosteroids, intravenous fluids, fluid balance, blood products and sedatives) until ICU discharge, death or day 30 in ICU, whichever came first. Follow-up was performed 90 days after discharge to record death or persistent organ dysfunction (defined as dependency on mechanical ventilation, renal replacement therapy or ongoing IV vasopressor use) (https://www.ualberta.ca/en/critical-care/media-library/documents/liberate-crf-v8.0_may_18_2023_clean.pdf).

### Sample Size

For the LIBERATE RCT, we calculated that a required sample size of 1,068 participants for the LIBERATE RCT would provide at least 80% power to detect a win ratio (WR) of 1.25 against a bull of no difference (i.e., WR = 1.0), with a two-sided alpha = 0.05. Using data from our systematic review and meta-analysis which showed ICU LOS estimates of 5.90 days (SD 4.89) in the midodrine treated cohort and 7.01 (SD 5.84) in the placebo cohort, ICU LOS was simulated as continuous, right-skewed log-normal outcome.[13] We conservatively assumed equal 28-mortality of 30% in both groups and applied the primary hierarchical endpoints of 28-day mortality followed by ICU LOS, using a 1-day threshold, which corresponds to the minimal clinically important difference for ICU LOS.[22] Across 1,000 simulated trials, the medical proportion of pairs remaining tied after the complete hierarchy was approximately 10%.

We retained a WR of 1.25 as the target clinically meaningful treatment effect. Using a target WR of 1.25, a 10% tie proportion, equal allocation, two-sided alpha of 0.05, and 80% power, the nonparametric large-sample WR calculations requires approximately 1,029 participants.

Allowing for a withdrawal rate of approximately 3%, this provides us with a recruitment target of 1,067 participants, rounded to 1,068 participants to maintain equal 1:1 allocation.

These implement the nonparametric large-sample approximation for the WR described by Wu and Ganju.[23] This approach does not require specification of parametric outcome distributions (i.e., log-normal or gamma for LOS). Instead, it requires specification of the target WR to be detected, the allocation ratio, and the anticipated proportion of ties, which together determine the variance of the log WR. We additionally performed simulation-based checks using mortality and ICU LOS distributions from our systematic review to determine operating characteristics, and the analytic calculation is distribution-free at the outcome levels; assumptions enter only through the anticipated WR and tie probability.

All statistics were calculated using the R package WR estimates and were conducted by our trial biostatistician (FZ).

### Statistical analysis

Analyses of the primary outcome was descriptive. Secondary outcomes involved summary measures obtained by aggregating the endpoints. No comparisons were conducted between groups as this multicenter pilot study was conducted to address trial objectives as outlined earlier; allocation primarily to assess recruitment rates and protocol adherence, retention rates, data completeness and safety. Results are reported herein as proportions and means with standard deviations, as appropriate.

### Management

To ensure protocol adherence and data quality, site initiation visits were held for research personnel at each site. In addition, the Coordinating Centre provided additional one on one training on request. A manual of procedures, training slides and website containing study documents (https://www.ualberta.ca/en/critical-care/research/liberate/index.html) were available to each research site. A newsletter containing frequently asked questions was published quarterly. This trial is Health Canada regulated (NOL obtained November 18, 2021). The Clinical Trials Office at the University of Alberta developed a detailed monitoring plan and conducted ongoing study monitoring of all study sites.

### Safety and adverse events

Reportable adverse events (AEs) were those considered to be related to study treatment or in the principal investigator’s clinical judgment, were not recognised events consistent with the participants underlying critical illness and/or chronic diseases and expected clinical course.

The reporting of adverse events followed the 5 recommendations for rational reporting of serious adverse events in investigator-initiated critical care trials of drugs in common use.[24] The adverse events to be reported included: clinically significant bradycardia (as per the judgment of the treating team); acute coronary syndrome (unstable angina, non-STEMI or STEMI); allergic event(s) (paresthesia, piloerection, dysuria, pruritis, chills, pain or rash); hypertension (sustained systolic measurement over 180mm Hg; bowel ischemia (clinically significant as per the judgement of the treating team); limb ischemia (clinically significant as per the judgement of the treating team); stroke (radiographical diagnosis); or other event considered to be related to study treatment or in the PI’s clinical judgement is not recognised as consistent with the subjects underlying critical illness and/or chronic disease and expected clinical course.

## Results

### Patient characteristics

Patients had a mean age of 61.7 (SD 15.0) and 80 (40.1%) were female. (Table 1) The mean Acute Physiology and Health Evaluation (APACHE II) score was 23.7 (SD 7.6) and the mean Sequential Organ Failure Assessment (SOFA) score at the time of study enrollment was 10.1 (SD 2.9). The mean Clinical Frailty Scale score was 4.1 (SD 1.6). Etiology of shock included: sepsis (63.4%, n=123), hypovolemia (12.4%, n=24), anaphylaxis (1.0%, n=2), cardiogenic (8.8%, n=17), neurogenic (6.2%, n=12), other cause (7.2%, n=14), and unknown (1.0%, n=2). The most frequently reported comorbid conditions at baseline included: diabetes mellitus (29.4%, n=57), respiratory insufficiency (18.6%, n=36), immune suppression (16.5%, n=32), coronary artery disease (13.4%, n=26), and congestive heart failure (11.3%, n=22).

**Table 1:** Baseline characteristics.

| Characteristics | n/N (%) |
| --- | --- |
| **Age, years** | **194/200 (97.0)** |
| Mean (SD) | 61.7 (15.0) |
| Median [IQR] | 64.5 [52, 72] |
| **Sex** | **194/200 (97.0)** |
| Female | 80 (40.1) |
| Male | 114 (59.0) |
| **Weight** | **194/200 (97.0)** |
| Mean (SD) | 82.3 (26.3) |
| Median [IQR] | 77.9 [63.8-95.9] |
| **Height** | **191/200 (95.5)** |
| Mean (SD) | 169.8 (13.4) |
| Median [IQR] | 170 [163-178] |
| **APACHEII** | **193/200 (97.0)** |
| Mean (SD) | 23.7 (7.6) |
| Median [IQR] | 23 [18-29] |
| **SOFA\*** | **180/200 (90.0)** |
| Mean (SD) | 10.1 (2.9) |
| Median [IQR] | 10 [8-12] |
| **CFS** | **191/200 (96.0)** |
| Mean (SD) | 4.1 (1.6) |
| Median [IQR] | 4 [3-5] |
| **Etiology of Shock** | **194/200 (97.0)** |
| Sepsis | 123 (63.4) |
| Hypovolemia | 24 (12.4) |
| Anaphylactic | 2 (1.0) |
| Cardiogenic | 17 (8.8) |
| Neurogenic | 12 (6.2) |
| Other | 14 (7.2) |
| Unknown | 2 (1.0) |
| **Any Comorbid Conditions?** |  |
| Coronary artery disease (Yes) | 26/194 (13.4) |
| Congestive heart failure (Yes) | 22/194 (11.3) |
| Respiratory insufficiency (Yes) | 36/194 (18.6) |
| Acute liver failure (Yes) | 8/194 (4.1) |
| Chronic liver failure (Yes) | 21/194 (10.8) |
| Chronic dialysis (Yes) | 6/194 (3.1) |
| Diabetes mellitus (Yes) | 57/194 (29.4) |
| Immune suppression (Yes) | 32/194 (16.5) |
| Leukemia/lymphoma (Yes) | 9/194 (4.6) |
| Metastatic cancer (Yes) | 13/194 (6.7) |

The SOFA score was implemented 9 months after the study started. Retrospective scores were not collected on the previous participants

Aside from IV vasopressors, ventilation was the most common reported intervention (88.1%, n=171), followed by sedation (59.8%, n=116), corticosteroids (34.5%, n=67), blood products (11.3%, n=22), and renal replacement therapy (5.7%, n=11). (Table 2) IV vasopressor and inotropic treatment included norepinephrine (99.5%, n=191), vasopressin (39.6%, n=76), epinephrine (3.1%, n=6), dobutamine (2.1%, n=4), and phenylephrine (2.1%, n=4) with approximately half the patients receiving one vasopressor (59.9%, n=115); others were receiving two or more vasopressors (40.2%, n=77) at time of study enrollment.

**Table 2:** ICU Interventions.

|  | **n/N (%)** |
| --- | --- |
| **Ventilatory Support** | **171/194 (88.1)** |
| Invasive | 131/171 (76.6) |
| Non-invasive | 4/171 (2.3) |
| High-flow oxygen | 8/171 (4.7) |
| Nasal cannula | 28/171 (16.4) |
| **Renal Replacement Therapy** | **11/194 (5.7)** |
| Intermittent Hemodialysis | 0/9 (0) |
| Prolonged Intermittent Renal Replacement Therapy | 0/9 (0) |
| Continuous Renal Replacement Therapy | 11/11 (100) |
| **Corticosteroids** | **67/194 (34.5)** |
| Hydrocortisone (IV) | 55/67 (82.1) |
| Hydrocortisone (PO) | 2/67 (3.0) |
| Dexamethasone (IV) | 7/67 (10.4) |
| Dexamethasone (PO) | 1/67 (1.5) |
| Methyl-prednisone | 1/67 (1.5) |
| Prednisone | 3/67 (4.5) |
| **Blood Products** | **22/194 (11.3)** |
| PRBC | 12/22 (54.5) |
| Albumin | 8/22 (36.4) |
| Platelets | 5/22 (22.7) |
| FFP | 1/22 (4.5) |
| Cryoprecipitate | -- |
| **Fluid balance Mean (SD)** | **1576.9 (2075.0)** |
| **IV Vasopressors or Inotropes** | **192/194 (99.0)** |
| Norepinephrine | 191/192 (99.5) |
| Vasopressin | 76/192 (39.6) |
| Epinephrine | 6/192 (3.1) |
| Dobutamine | 4/192 (2.1) |
| Phenylephrine | 4/192 (2.1) |
| **Number of vasopressors at the time of study enrollment** |  |
| 1 | 115/192 (59.9) |
| 2 | 65/192 (33.9) |
| 3 | 12/192 (6.3) |
| **Sedation** | **116/194 (59.8)** |
| Propofol | 100/116 (86.2) |
| Hydromorphone | 22/116 (19.0) |
| Fentanyl | 22/116 (19.0) |
| Midazolam | 6/116 (5.2) |
| Ketamine | 3/116 (2.6) |
| Morphine | 1/116 (0.9) |

### Primary Outcomes

#### Recruitment rate

The primary feasibility outcomes are outlined in Table 3. A total of 200 patients were enrolled over a 29-month period with an average overall recruitment rate of 7.0 [range 1-20] patients per month and a per-site recruitment rate of 1.4 (range 0.3-3.0) patients per month.

**Table 3:**
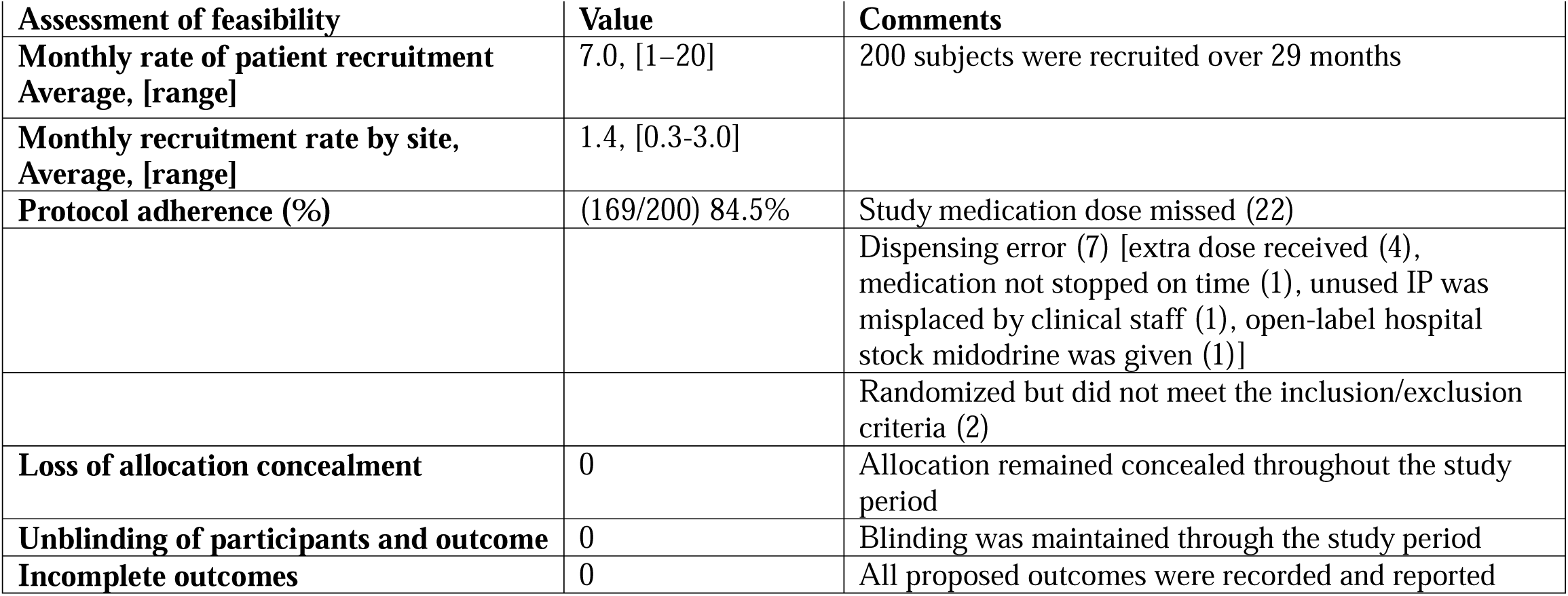
Feasibility outcomes.

#### Screening and enrollment characteristics

Screening statistics were reported from 5 of the 9 sites (n=85, 42.5%); the other 4 sites did not collect screening statistics. For those sites reporting, 705 patients were screened and 620 were excluded. The 3 most common causes for non-enrollment were: a contraindication to enteral medication (n=199, 31.4%); over 24 hours from peak vasopressor dose (n=141, 22.2%); and expected death or withdrawal of treatment (n=98, 15.4%) (Figure 1).

**Figure 1.**
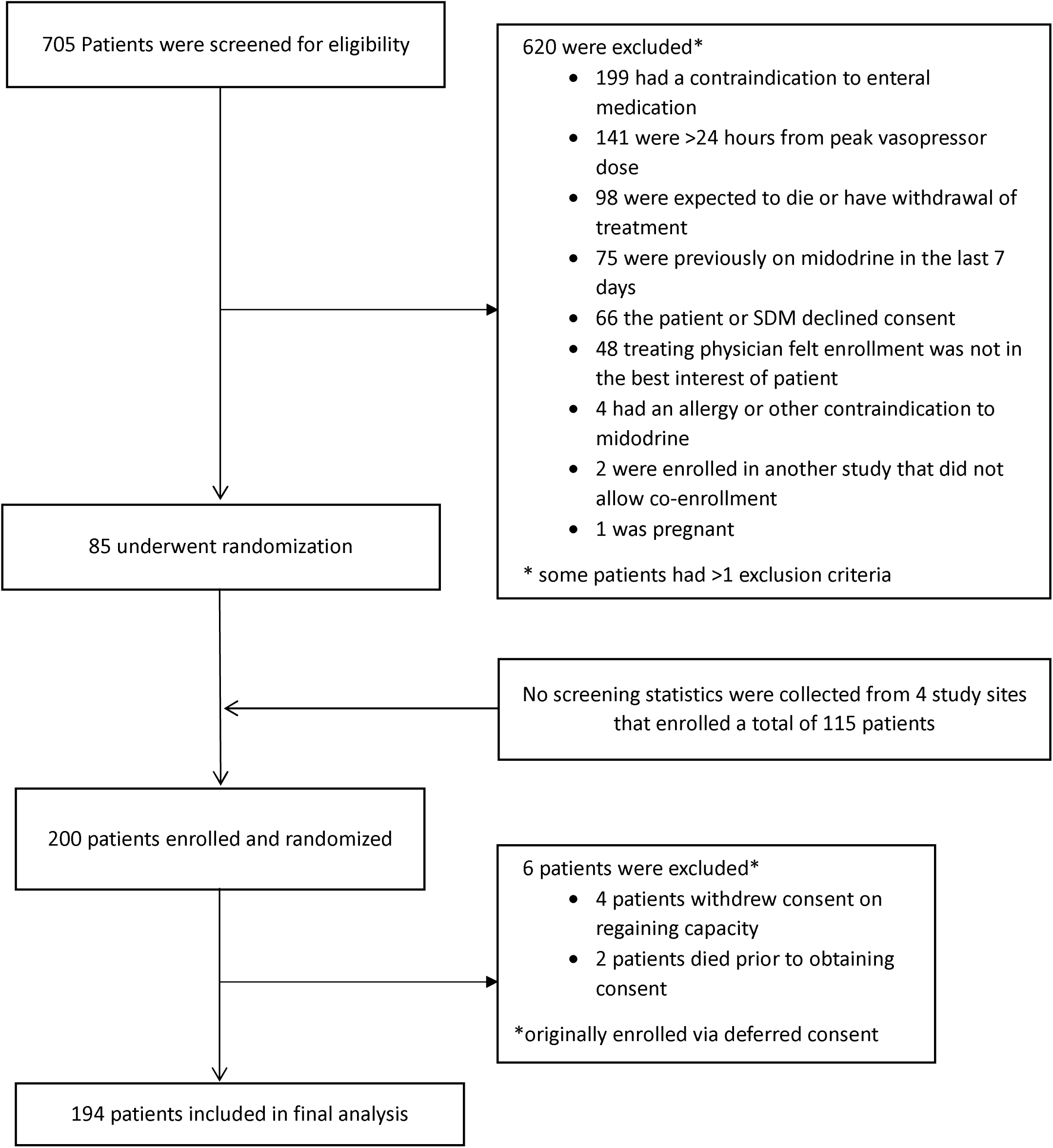
Participant Flow Diagram. Screening characteristics are only available from 5 out of 9 sites that maintained screening logs.

Overall, consent was obtained from 39 patients (19.5%), while an SDM provided consent for 78 (39.0%) of the patients and 83 (41.5%) of patients were enrolled under the deferred consent model.

#### Protocol adherence, retention and data completeness

Protocol adherence was 84.5%, with study medication dose missed/dispensing errors being the most common deviation (14.9%, 29 occurrences) followed by randomized but did not meet inclusion/exclusion criteria (1.0%, 2 occurrences). There were no reports of unblinding of treatment allocation by the study sites. The retention rate was 97.0% with 4 patients withdrawing consent once they regained capacity and 2 patients who died before they could be approached for a regained capacity consent, for whom the local REB did not provide a waiver of consent. Data completeness was high with an average completion rate of 96.0%.

#### Adverse events

Adverse events in two participants (1.0%) were reported. One patient experienced hypertension (defined as sustained systolic measurement over 180mm Hg) and one patient’s heart rate decreased to 41 beats per minute (clinically significant bradycardia as per the judgement of the treating team). Both events were assessed as moderate in severity, probable for a causal relationship between the study treatment and adverse event, and in both cases, the study treatment was permanently discontinued. Both patients recovered from the adverse event with no sequelae. There were no other serious adverse events reported.

### Secondary outcomes

The secondary clinical outcomes are outlined in Table 4, reported in aggregate. The mean length of IV vasopressor support was 10.3 (SD 5.1 hours) and IV vasopressors were reinitiated over 24 hours after cessation in 18.0% (n=35) of patients. Mean ICU and hospital lengths of stay were 10.6 days (SD: 14.7) and 25.8 days (SD: 37.3), respectively. ICU death occurred in 15.5% (n=30) of patients and hospital death in 23.6% (n=51). 2 patient (1.0%) was readmitted to ICU within 48 hours of discharge.

**Table 4:**
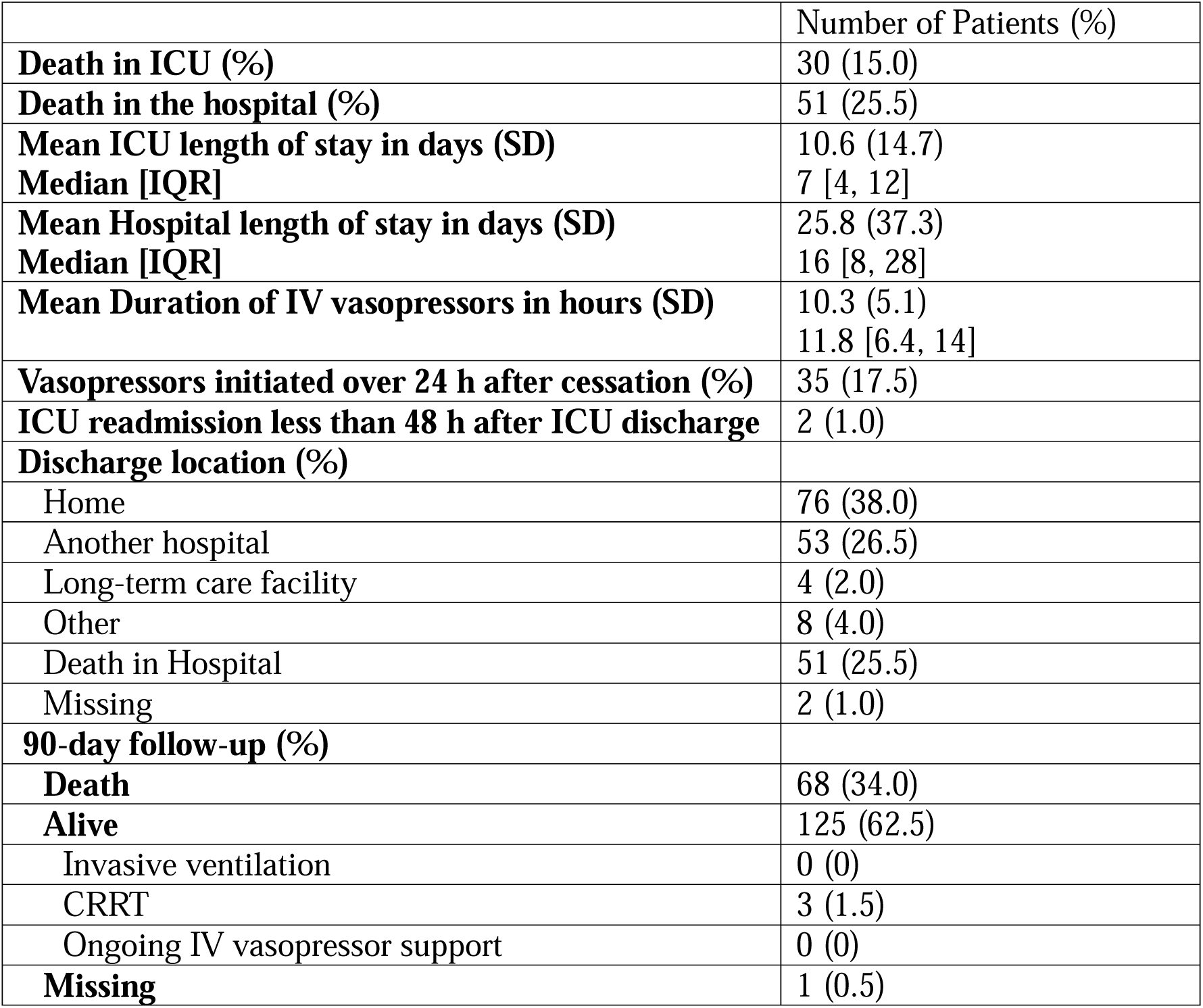
Clinical outcomes.

Death at 90 days was 35.1% (n=68), and only 3 patients (2.4%) required ongoing renal replacement therapy at 90 days.

### Trial oversight

A Management Committee comprised of the Principal Investigator and Project Manager acted as the Coordinating Centre and oversaw the study set up and day-to-day activities. The Management Committee met weekly to manage development and execution of the study and ensure appropriate adherence to the study protocol, and was responsible for reporting to and updating the Steering Committee and the Data Safety Management Committee of all study proceedings.

The Steering Committee is a multidisciplinary body consisting of research clinicians, a patient-partner, an epidemiologist and a health economics expert. The Steering Committee meets quarterly and provides input on all matters related to the research and protocol agenda and is also responsible for local endorsement of the program of work.

The Data Safety Monitoring Committee (DSMC) is an independent body that safeguards the interests of study participants by assessing the safety of study procedures and provides recommendations about continuing or stopping the study based on safety considerations. Every 6 months the DSMC assessed data quality; monitored protocol compliance and participant recruitment, accrual and retention; reviewed adverse event and serious adverse event data; and assessed any external data that may impact the safety of the participants or the ethics of the study.

The LIBERATE trial is a Canadian Critical Care Trials (CCCTG) endorsed research program. The CCCTG is a pan-Canadian partnership of multi-disciplinary, interprofessional researchers dedicated to the pursuit of excellence and advancement of critical care research in Canada.[25]

## Discussion

The LIBERATE internal randomized controlled trial established the feasibility of conducting the LIBERATE RCT across multiple centers in multiple jurisdictions in a variety of different settings. This confirmed the feasibility results from our previous single-centre feasibility study across additional centres.[20] We determined a mean recruitment rate of 1.4 patients/month/site (range 0.3-3), a screening and enrollment procedure that achieved recruitment targets, adequate protocol adherence (84.5%), high study retention (97.0%) and completeness of data (96.0%), and only 2 reportable adverse events. Further, we demonstrated that the LIBERATE internal randomized controlled trial study was feasible at both academic and community sites, having established a mechanism to provide IP in a blinded manner to sites without a dedicated research pharmacy.

IV vasopressor therapy for dependent shock is one of the most common forms of physiological support delivered in ICUs.[26] This occurs across all centers. Further, ICU capacity strain is not limited to a specific type of ICU, and the potential use of midodrine to decrease ICU length of stay is applicable to all ICUs. Having established feasibility at both academic and community sites, the LIBERATE study will ensure engagement with community sites in order to provide equitable access to clinical research and build research capacity in the ICU research community. This is a key goal of the CCCTG through the Canadian Community ICU Research Network.[27–29] To that end, the study protocol was developed according to the Pragmatic-Explanatory Continuum Indicator Summary (PRECIS-2) framework for pragmatic trials.[30] This pragmatic protocol, which evaluates an inexpensive, readily available therapy, was designed to maximize acceptability at non-traditional research sites, such as community hospitals. One item that emerged was the inconsistencies among sites in collecting enrollment data and other complexities and variabilities in recruitment dynamics across different ICUs. This is consistent with findings from previous critical care RCTs.[31] This variability highlights the need to protocolize the recording of recruitment dynamics and enrollment data for the LIBERATE trial. The LIBERATE internal randomized controlled trial is the largest RCT evaluating the use of midodrine in critically ill patients to date. Further, it is the RCT with the highest number of participating sites. The MIDAS study by Santer *et al*, is next largest. However, this study evaluated 132 patients across 3 centers and required 7 years to recruit this sample size. Additionally, with an extensive list of exclusion criteria that may not truly represent critically ill patients requiring IV vasopressor, may have led to a possible selection bias. It also focused on a non-patient centered outcome (i.e., time to IV vasopressor discontinuation) and did not include a subgroup analysis. Finally, in our recent systematic review, our time-sequential analysis revealed a minimal sample size of 1124 patients to determine the effects of midodrine on ICU LOS. While the LIBERATE internal randomized controlled trial will add to the published literature, we will not yet achieve the minimal sample size necessary, as determined by our systematic review.

However, the LIBERATE internal randomized controlled trial has established feasibility for the LIBERATE study, which aims to recruit 1,068 participants across 25 centers, which will definitely determine the utility of adjunctive midodrine therapy for critically ill patients with IV vasopressor-dependent shock.

Limitations of this study include heterogeneity of recruitment rates between study sites (range 0.3-3.0), although a sufficient number of patients were enrolled overall to meet our recruitment objectives. Reasons provided by low-enrolling sites were mainly due to internal resource issues (i.e., inadequate number of research staff, high turnover rate of research staff). The range of recruitment rates will inform our study center to assist sites in achieving adequate recruitment into the LIBERATE study. We have also identified barriers and facilitators for recruitment, and will communicate these with sites. Other important limitations were protocol deviations due to errors in medication dispensing, administration and storage, and that screening logs were not maintained at all study sites. Potential strategies would include, 1) offering additional one on one training sessions for new or existing research staff, and 2) discussing strategies or providing additional tools (i.e., IP discontinuation reminder cards) to mitigate medication errors. In addition, increasing the uptake of screening logs could be achieved by being more prescriptive in the expectations of the study sites or by providing additional financial compensation if a resource issue is identified.

## Conclusions

The utility of midodrine in critically ill patients with IV vasopressor shock is uncertain. The LIBERATE internal randomized controlled trial is a vital first step to fulfilling this critical knowledge gap and informing practice.

## Data Availability

Data produced in the present study are available upon reasonable request to the authors.
However, group clinical outcome data remain blinded to the authors as this study is the vanguard phase of the LIBERATE trial.

## DECLARATIONS

### Consent for publication

We confirm that this manuscript has been read and approved by all named authors and that there are no other persons who satisfied the criteria for authorship but are not listed. We further confirm that the order of authors listed in the manuscript has been approved by all.

We confirm we have given due consideration to the protection of intellectual property associated with this work and that there are no impediments to publication, including the timing of publication with respect to intellectual property. In doing so we confirm that we have followed the regulation of our institutions concerning intellectual property.

### Ethics approval and consent to participate

We further confirm any aspect of the work covered in this manuscript that has involved human patients has been conducted with the ethical approval of all relevant bodies and that such approvals are acknowledged within the manuscript. We understand that the Corresponding Author is the sole contact for the Editorial process (including Editorial Manager and direct communications with the office). He is responsible for communicating with the other authors about progress, submissions of revisions and final approval of proofs. We confirm that we have provided a current, correct email address which is accessible by the Corresponding Author.

### Availability of data and materials

Not applicable.

### Contributors

DO contributed to study protocol development and drafting of the manuscript, FH, DJC, DD, SS, JT, JS, VL and FZ contributed to study protocol development and critical revision of the manuscript, SMB and OGR conceived the study, developed the protocol and contributed to development and drafting of the manuscript. All of the authors approved the final version to be published.

## Acknowledgements

SB is supported by a Canada Research Chair in Critical Care Outcomes and Systems Evaluation. DJC is supported by a Canada Research Chair in Knowledge Translation in Critical Care.

## LIBERATE multi-centre pilot trial clinical collaborators

Dr. Janek Senaratne, Anushka Jayasekara, Bernadette Fernando, Isabel Kwek, Larissa Fedor-Turchenek, Grey Nuns Community Hospital, Edmonton, AB

Dr. Maureen Meade, Lori Hand, Hamilton Health Sciences, Hamilton General Hospital, Hamilton, ON

Dr. Erika Macintyre, Daniel Ofosu, Misericordia Community Hospital, Edmonton, AB

Dr. Sangeeta Mehta, Sumesh Shah, Mount Sinai Hospital, Toronto, ON

Dr. Jocelyn Slemko, Sargun Sokhi, Sturgeon Community Hospital, Edmonton, AB

Dr. Deborah Cook, France Clarke, St Joseph’s Healthcare, Hamilton, ON

Dr. Jennifer Tsang, Anna Kim, Emily Baker, Lisa Patterson, Niagara Health, St Catharines, St Catharines, ON

Dr Victor Dong, Ish Bains, Rockyview General Hospital, Calgary, AB

Dr. Stephanie Sibley, Miranda Hunt, Tracy Boyd, Kingston Health Sciences Centre, Kingston, ON

Dr. Oleksa Rewa, Caylin Chadwick, Nadia Baig, Lily Guan, University of Alberta Hospital, Edmonton, AB

## List of Abbreviations

IV: intravenous
RCT: randomized controlled trial
LOS: length of stay
SDM: substitute decision maker
IP: investigational product
APACHE: Acute Physiology and Chronic Health Evaluation
SOFA: Sequential Organ Failure Assessment
STEMI: ST-elevation myocardial infarction

